# Baseline QRS Duration Associates with Cardiac Recovery in Patients with Continuous-Flow Left Ventricular Assist Device Implantation

**DOI:** 10.1101/2021.09.21.21263287

**Authors:** Muhammad S. Khan, Christos P. Kyriakopoulos, Iosif Taleb, Elizabeth Dranow, Monte Scott, Ravi Ranjan, Michael Yin, Eleni Tseliou, Rami Alharethi, William Caine, Robin M. Shaw, Craig H. Selzman, Stavros G. Drakos, Derek J. Dosdall

**Affiliations:** Nora Eccles Harrison Cardiovascular Research and Training Institute, The University of Utah, Salt Lake City, UT; Division of Cardiovascular Medicine, Department of Internal Medicine, University of Utah Health & School of Medicine, Salt Lake City, UT; Department of Biomedical Engineering, The University of Utah, Salt Lake City, UT; Cardiovascular Department, Intermountain Medical Center, Salt Lake City, UT; Division of Cardiothoracic Surgery, Department of Surgery, University of Utah Health & School of Medicine, Salt Lake City, UT

**Keywords:** QRS duration, LVAD, cardiac recovery, LVEF, LVEDD, electrical remodeling

## Abstract

**BACKGROUND:** In chronic heart failure (HF) patients supported with continuous-flow left ventricular assist device (CF-LVAD), we aimed to assess the clinical association of baseline QRS duration (QRSd) with post-LVAD cardiac recovery, and its correlation with pre- to post-LVAD change in left ventricular ejection fraction (LVEF) and left ventricular end-diastolic diameter (LVEDD).

**METHODS:** Chronic HF patients (n=402) undergoing CF-LVAD implantation were prospectively enrolled, at one of the centers comprising the U.T.A.H. (Utah Transplant Affiliated Hospitals) consortium. After excluding patients with acute HF etiologies, hypertrophic or infiltrative cardiomyopathy, and/or inadequate post-LVAD follow up (<3 months), 315 patients were included in the study. Cardiac recovery was defined as LVEF ≥40% and LVEDD <6 cm within 12 months post-LVAD implantation. Patients fulfilling this condition were termed as responders (R) and results were compared with non-responders (NR).

**RESULTS:** Thirty-five patients (11%) achieved ‘R’ criteria, and exhibited a 15% shorter QRSd compared to ‘NR’ (123±37 ms vs 145±36 ms; p<0.001). A univariate analysis identified association of baseline QRSd with post-LVAD cardiac recovery (OR:0.986, 95% CI:0.976-0.996, p<0.001). In a multivariate logistic regression model, after adjusting for duration of HF (OR:0.990, 95% CI:0.983-0.997, p=0.006) and gender (OR:0.388, 95% CI:0.160-0.943, p=0.037), pre-LVAD QRSd exhibited a significant association with post-LVAD cardiac structural and functional improvement (OR:0.987, 95% CI:0.977-0.998, p=0.027) and the predictive model showed a c-statistic of 0.73 with p<0.001. The correlations for baseline QRSd with pre- to post-LVAD change in LVEF and LVEDD were also investigated in ‘R’ and ‘NR’ groups.

**CONCLUSION:** Chronic advanced HF patients with a shorter baseline QRSd exhibit an increased potential for cardiac recovery after LVAD support.

## INTRODUCTION

In patients with advanced heart failure (HF) refractory to medical therapy, continuous-flow (CF) left ventricular (LV) assist devices (LVADs) have been used as a bridge to transplantation,^1,2^ as destination therapy,^3^ as a bridge to transplant candidacy, and/or as a bridge to recovery.^4,5^ The number of LVAD implantations has continued to grow in the US in comparison with the number of heart transplantations.^6,7^ While left ventricular ejection fraction (LVEF) during mechanical unloading is used to identify patients achieving cardiac recovery, it has showed no predictive value prior to LVAD implantation.^8^ LV torsion has been found to play a pivotal role in facilitating the homogenous distribution of myocardial forces during systole.^9^ Clinical studies in chronic HF patients have associated LV rotational dynamics with the degree of remodeling and the extension of myocardial fibrosis.^10,11^ In addition, LV global longitudinal strain has been previously studied and correlated with the extent of myocardial fibrosis in patients with advanced HF.^12,13^

Previous studies have focused on a prolonged QRS duration (QRSd) that appears common in patients with reduced LVEF and were hospitalized for HF management.^14–16^ Further, the impact of LVAD unloading on the electrical properties (QRS, QT and QTc duration) of the failing heart has also been reported.^17^ In this study, we sought to examine whether baseline QRSd associates with post-LVAD cardiac recovery in chronic heart failure (CHF) patients undergoing LVAD implantation. We further demonstrate the correlations for baseline QRSd with pre- and post-LVAD LVEF and LVEDD in LVAD patients, and compared the data of those who showed a successful cardiac recovery with those who did not show recovery within 12 months of LVAD support. Finally, in addition to univariate and bivariate analyses, a multivariate logistic regression model is reported including other clinical parameters to find whether the baseline QRSd is independently associated with post-LVAD cardiac recovery.

## METHODS

### Study Population

Advanced cardiomyopathy patients (n=402) undergoing LVAD implantation at one of the institutions comprising the Utah Transplant Affiliated Hospitals (U.T.A.H.) Cardiac Transplant Program (i.e. University of Utah Health, Intermountain Medical Center, and George E. Wahlen Veterans Affairs Medical Center) were prospectively enrolled. The study was approved by the Institute Review Board (IRB) -The University of Utah, Salt Lake City, UT 84112. Ethical approval was given, and the patients were prospectively enrolled. Informed consent was obtained under the IRB 30622 - “Effects of Mechanical Unloading on Myocardial Function and Structure in Humans study.” Acute HF etiologies, hypertrophic or infiltrative cardiomyopathy, baseline LVEF ≥40%, and inadequate post-LVAD follow up (<3 months) were the exclusion criteria. Our final study cohort included a total of 315 patients [56±15 years old, 267 (85%) male] as shown in **Fig. 1**. The patients’ long-term medications regimen before LVAD implantation included ß-blockers, angiotensin-converting enzyme inhibitors (ACEI), angiotensin II receptor blockers (ARB), aldosterone antagonists, and diuretics. About 71% patients were NYHA class IV. Implanted devices were HeartMate II™ (n=121), HeartMate 3™ (n=17), HeartWare™ (n=156) and others (n=21).

**Figure 1.**
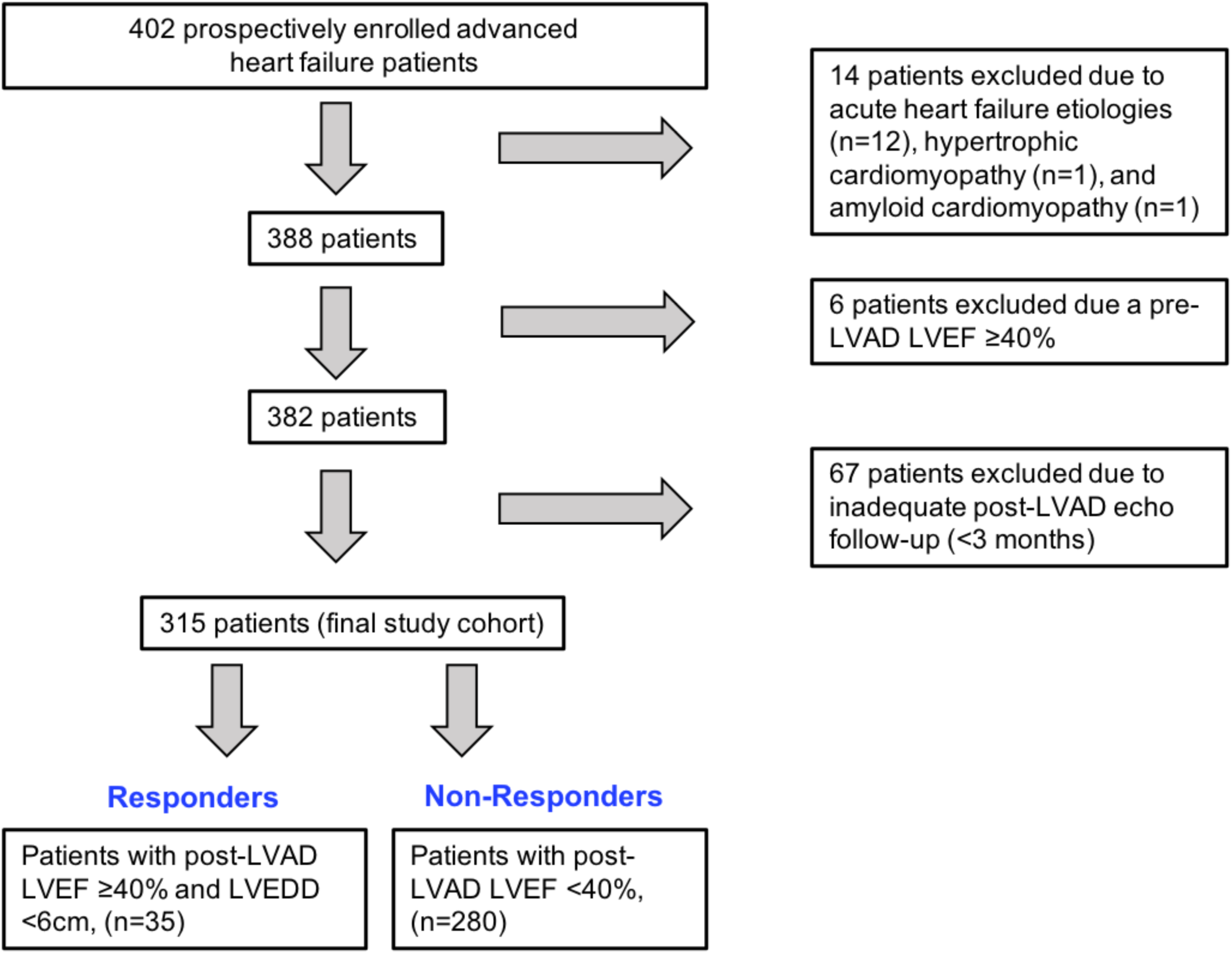
Flowchart description of advanced HF patients undergoing LVAD implantation and included in the study.

### LVAD-induced Cardiac Recovery Definition

LVAD-induced cardiac recovery was defined as an LVEF ≥40% and LVEDD <6 cm within 12 months post-LVAD implantation. Patients fulfilling the above criteria were termed responders (R) (n=35) with their counterparts not achieving significant cardiac structural and functional improvement following LVAD implantation, constituting the non-responders (NR) group (n=280).

### Data Collection

Demographic, medication, laboratory, hemodynamic and electrocardiographic data were collected within 1 week prior to LVAD implantation. Transthoracic echocardiograms were performed within 2 weeks preceding LVAD implant, and then serially at months 1, 2, 3, 4, 6, 9, and 12 after implantation, using a protocol developed and tested at the Utah Cardiac Recovery Program.^18^ Complete 2-dimensional, M-mode, and Doppler images were recorded from standard views in accordance with current American Society of Echocardiography guidelines.^19^ Last available reported LVEF and LVEDD values within 1-year post-LVAD implant were used to assess cardiac recovery.

### Statistical Analysis

Continuous variables are expressed as mean±standard deviation and were compared using unpaired t-test. Categorical variables are expressed as counts and percentages and were compared using chi-square test. Covariates remaining at the last step were included in the multivariable model to evaluate baseline QRS as an independent predictor for post-LVAD cardiac recovery. The presence of collinearity among candidate covariates was assessed with the variance inflation factor diagnostic.^20^ A bootstrap inclusion fraction (BIF) was calculated for each potential predictor, defined as the percentage of time that each variable would be retained in the model as a significant predictor in 1000 bootstrap resamples, in which the backwards elimination variable selection is repeated.^21^ Variables with BIFs <50% were dropped from the model as unreliable, as these would not likely remain significant predictors in external data sets. A p-value<0.10 was used to screen covariates for inclusion in the multivariate analysis. Receiver-operator-characteristics curve analysis was performed to determine the accuracy of pre-LVAD QRSd along with other potential variables to predict post-LVAD cardiac recovery. Based on a previously reported method,^22^ a truncation approach was also applied to overcome the missing data, equipment limit resolution and to remove outliers in both the ‘R’ and ‘NR’ groups. All significance tests were 2-tailed, and p<0.05 was considered statistically significant. Statistical analyses were performed using Stata 16.0.^23^

## RESULTS

Among 315 chronic HF patients included in the analysis, 35 patients achieved cardiac recovery while on LVAD support (R). We summarized the baseline clinical characteristics, medications, laboratory results, hemodynamic and echocardiography parameters of ‘R’ and ‘NR’ in **Table 1**. Following sections will elaborate on the differences between the two groups of LVAD patients.

**Table 1.**
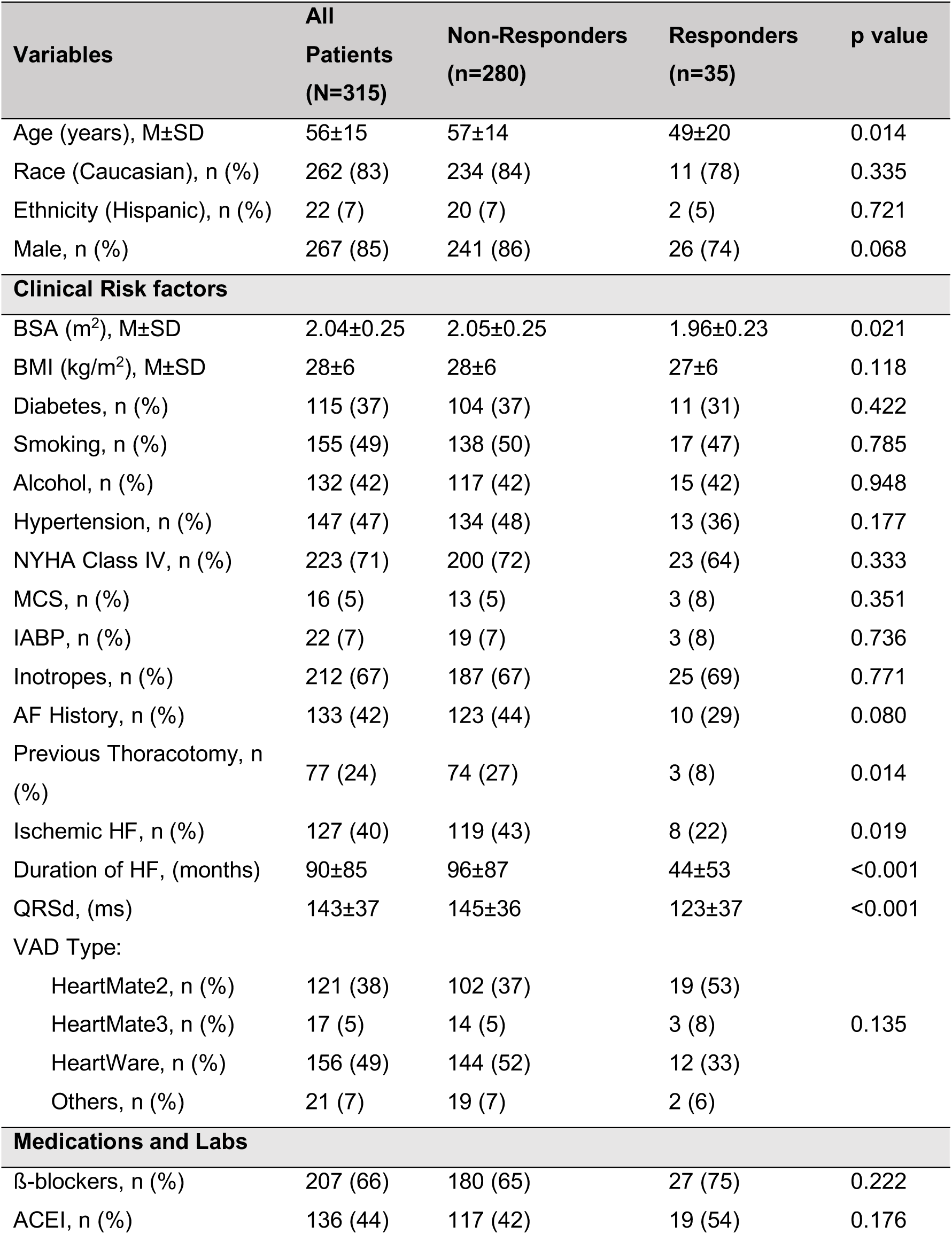
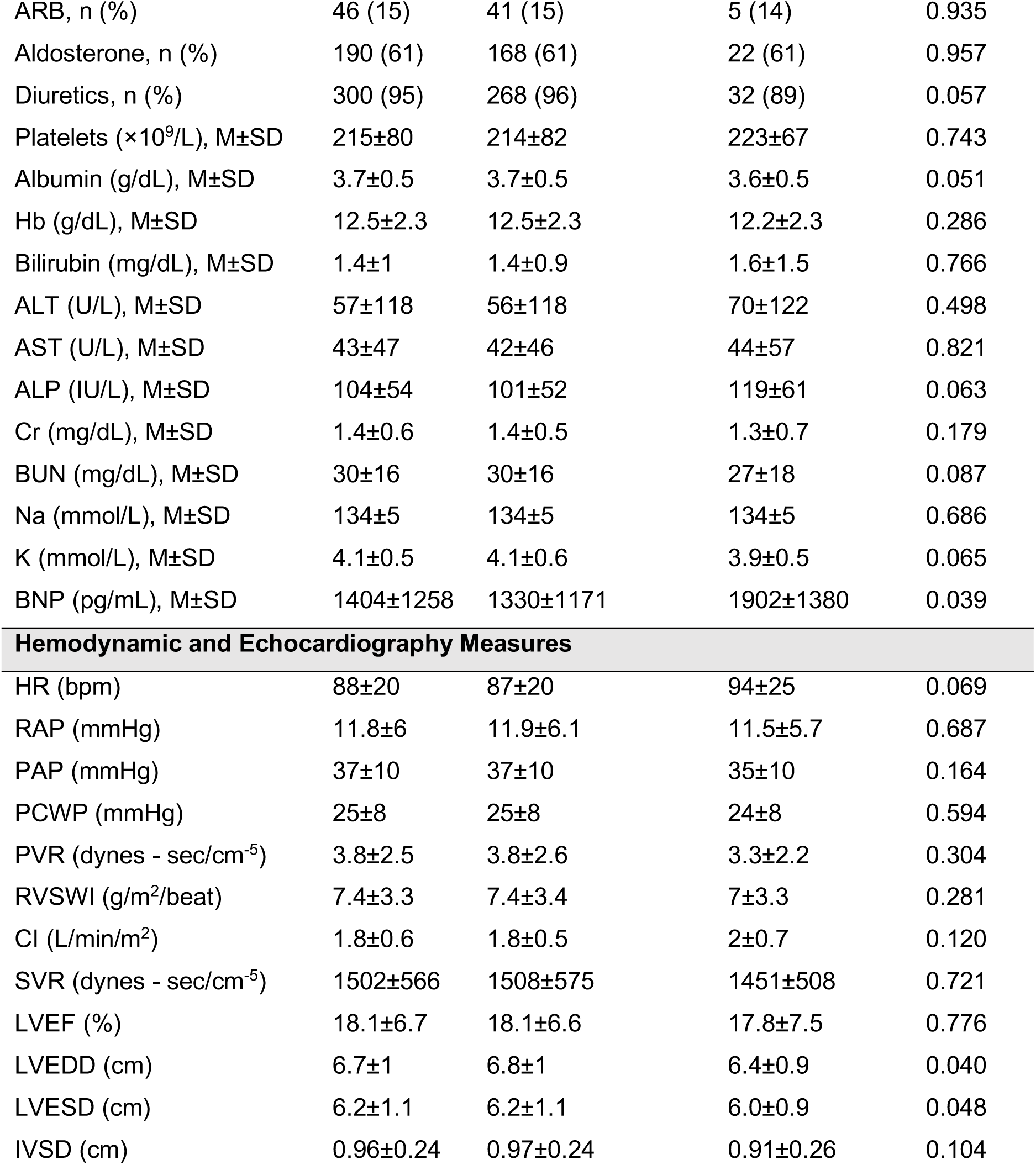
Demographics, baseline clinical characteristics, medications, labs, hemodynamic and echocardiography measures of responders and non-responders.

| Variables | All Patients<br>(N=315) | Non-Responders<br>(n=280) | Responders<br>(n=35) | p value |
| --- | --- | --- | --- | --- |
| Age (years), M±SD | 56±15 | 57±14 | 49±20 | 0.014 |
| Race (Caucasian), n (%) | 262 (83) | 234 (84) | 11 (78) | 0.335 |
| Ethnicity (Hispanic), n (%) | 22 (7) | 20 (7) | 2 (5) | 0.721 |
| Male, n (%) | 267 (85) | 241 (86) | 26 (74) | 0.068 |
| Clinical Risk factors |  |  |  |  |
| BSA (m<sup>2</sup>), M±SD | 2.04±0.25 | 2.05±0.25 | 1.96±0.23 | 0.021 |
| BMI (kg/m<sup>2</sup>), M±SD | 28±6 | 28±6 | 27±6 | 0.118 |
| Diabetes, n (%) | 115 (37) | 104 (37) | 11 (31) | 0.422 |
| Smoking, n (%) | 155 (49) | 138 (50) | 17 (47) | 0.785 |
| Alcohol, n (%) | 132 (42) | 117 (42) | 15 (42) | 0.948 |
| Hypertension, n (%) | 147 (47) | 134 (48) | 13 (36) | 0.177 |
| NYHA Class IV, n (%) | 223 (71) | 200 (72) | 23 (64) | 0.333 |
| MCS, n (%) | 16 (5) | 13 (5) | 3 (8) | 0.351 |
| IABP, n (%) | 22 (7) | 19 (7) | 3 (8) | 0.736 |
| Inotropes, n (%) | 212 (67) | 187 (67) | 25 (69) | 0.771 |
| AF History, n (%) | 133 (42) | 123 (44) | 10 (29) | 0.080 |
| Previous Thoracotomy, n (%) | 77 (24) | 74 (27) | 3 (8) | 0.014 |
| Ischemic HF, n (%) | 127 (40) | 119 (43) | 8 (22) | 0.019 |
| Duration of HF, (months) | 90±85 | 96±87 | 44±53 | <0.001 |
| QRSd, (ms) | 143±37 | 145±36 | 123±37 | <0.001 |
| VAD Type: |  |  |  |  |
| HeartMate2, n (%) | 121 (38) | 102 (37) | 19 (53) | 0.135 |
| HeartMate3, n (%) | 17 (5) | 14 (5) | 3 (8) |  |
| HeartWare, n (%) | 156 (49) | 144 (52) | 12 (33) |  |
| Others, n (%) | 21 (7) | 19 (7) | 2 (6) |  |
| Medications and Labs |  |  |  |  |
| β-blockers, n (%) | 207 (66) | 180 (65) | 27 (75) | 0.222 |
| ACEI, n (%) | 136 (44) | 117 (42) | 19 (54) | 0.176 |
| ARB, n (%) | 46 (15) | 41 (15) | 5 (14) | 0.935 |
| Aldosterone, n (%) | 190 (61) | 168 (61) | 22 (61) | 0.957 |
| Diuretics, n (%) | 300 (95) | 268 (96) | 32 (89) | 0.057 |
| Platelets ($\times 10^9/L$), M±SD | 215±80 | 214±82 | 223±67 | 0.743 |
| Albumin (g/dL), M±SD | 3.7±0.5 | 3.7±0.5 | 3.6±0.5 | 0.051 |
| Hb (g/dL), M±SD | 12.5±2.3 | 12.5±2.3 | 12.2±2.3 | 0.286 |
| Bilirubin (mg/dL), M±SD | 1.4±1 | 1.4±0.9 | 1.6±1.5 | 0.766 |
| ALT (U/L), M±SD | 57±118 | 56±118 | 70±122 | 0.498 |
| AST (U/L), M±SD | 43±47 | 42±46 | 44±57 | 0.821 |
| ALP (IU/L), M±SD | 104±54 | 101±52 | 119±61 | 0.063 |
| Cr (mg/dL), M±SD | 1.4±0.6 | 1.4±0.5 | 1.3±0.7 | 0.179 |
| BUN (mg/dL), M±SD | 30±16 | 30±16 | 27±18 | 0.087 |
| Na (mmol/L), M±SD | 134±5 | 134±5 | 134±5 | 0.686 |
| K (mmol/L), M±SD | 4.1±0.5 | 4.1±0.6 | 3.9±0.5 | 0.065 |
| BNP (pg/mL), M±SD | 1404±1258 | 1330±1171 | 1902±1380 | 0.039 |
| Hemodynamic and Echocardiography Measures |  |  |  |  |
| HR (bpm) | 88±20 | 87±20 | 94±25 | 0.069 |
| RAP (mmHg) | 11.8±6 | 11.9±6.1 | 11.5±5.7 | 0.687 |
| PAP (mmHg) | 37±10 | 37±10 | 35±10 | 0.164 |
| PCWP (mmHg) | 25±8 | 25±8 | 24±8 | 0.594 |
| PVR (dynes - sec/cm<sup>-5</sup>) | 3.8±2.5 | 3.8±2.6 | 3.3±2.2 | 0.304 |
| RVSWI (g/m<sup>2</sup>/beat) | 7.4±3.3 | 7.4±3.4 | 7±3.3 | 0.281 |
| CI (L/min/m<sup>2</sup>) | 1.8±0.6 | 1.8±0.5 | 2±0.7 | 0.120 |
| SVR (dynes - sec/cm<sup>-5</sup>) | 1502±566 | 1508±575 | 1451±508 | 0.721 |
| LVEF (%) | 18.1±6.7 | 18.1±6.6 | 17.8±7.5 | 0.776 |
| LVEDD (cm) | 6.7±1 | 6.8±1 | 6.4±0.9 | 0.040 |
| LVESD (cm) | 6.2±1.1 | 6.2±1.1 | 6.0±0.9 | 0.048 |
| IVSD (cm) | 0.96±0.24 | 0.97±0.24 | 0.91±0.26 | 0.104 |

### Baseline Characteristics of LVAD Patients with Cardiac Recovery

Among many baseline and clinical parameters outlined in **Table 1**, age, body surface area (BSA), previous thoracotomy, ischemic HF etiology and pre-LVAD HF duration were significantly different in the ‘R’ and ‘NR’ groups. For example, the group of patients who did not respond to LVAD support were older than those who responded within 12 months of LVAD support (57±14 vs. 49±20 years, p=0.014), and also pre-LVAD BSA of ‘NR’ group was higher (2.05±0.25 vs. 1.96±0.23 m^2^, p=0.021) in comparison to the ‘R’ group. Further, in comparison to the ‘R’ group, patients in the ‘NR’ group had a significantly longer HF duration (96±87 vs. 44±53 months, p<0.001) with a history of previous thoracotomy (p=0.014) and an ischemic HF etiology (p=0.019) as detailed in **Table 1**.

Regarding laboratory results, baseline B-type natriuretic peptide (BNP) level in the ‘R’ group was significantly higher in comparison to the ‘NR’ group (1902±1380 pg/mL vs. 1330±1171, p=0.039). Finally, pre-LVAD LVEDD and LVESD of ‘R’ group LVAD patients were significantly lower as compared to ‘NR’ group and reported as 6.4±0.9 vs. 6.8±1.0 cm (p=0.040) and 6.0±0.9 vs. 6.2±1.1 cm (p=0.048), respectively. Details of other significant / non-significant clinical, laboratory, hemodynamic and echocardiographic parameters are listed in **Table 1**.

### Baseline QRSd in LVAD Patients

The mean baseline QRSd of the total study population was 143±37 ms. Pre-LVAD QRSd in the ‘R’ group was 14.5% shorter than the duration reported in the ‘NR’ group (123±37 ms vs. 145±36 ms, respectively, p<0.001), as shown in **Fig. 2**. Interestingly, LVEF did not differ significantly between the ‘R’ and ‘NR’ groups before LVAD implantation (17.8±7.5 vs. 18.1±6.6 %, p=0.776). Based on univariate logistic regression (**Table 2**), pre-LVAD QRSd shows a significant association with post-LVAD cardiac recovery in LVAD patients (OR: 0.983, 95% CI: 0.972-0.993, p<0.001).

**Figure 2.**
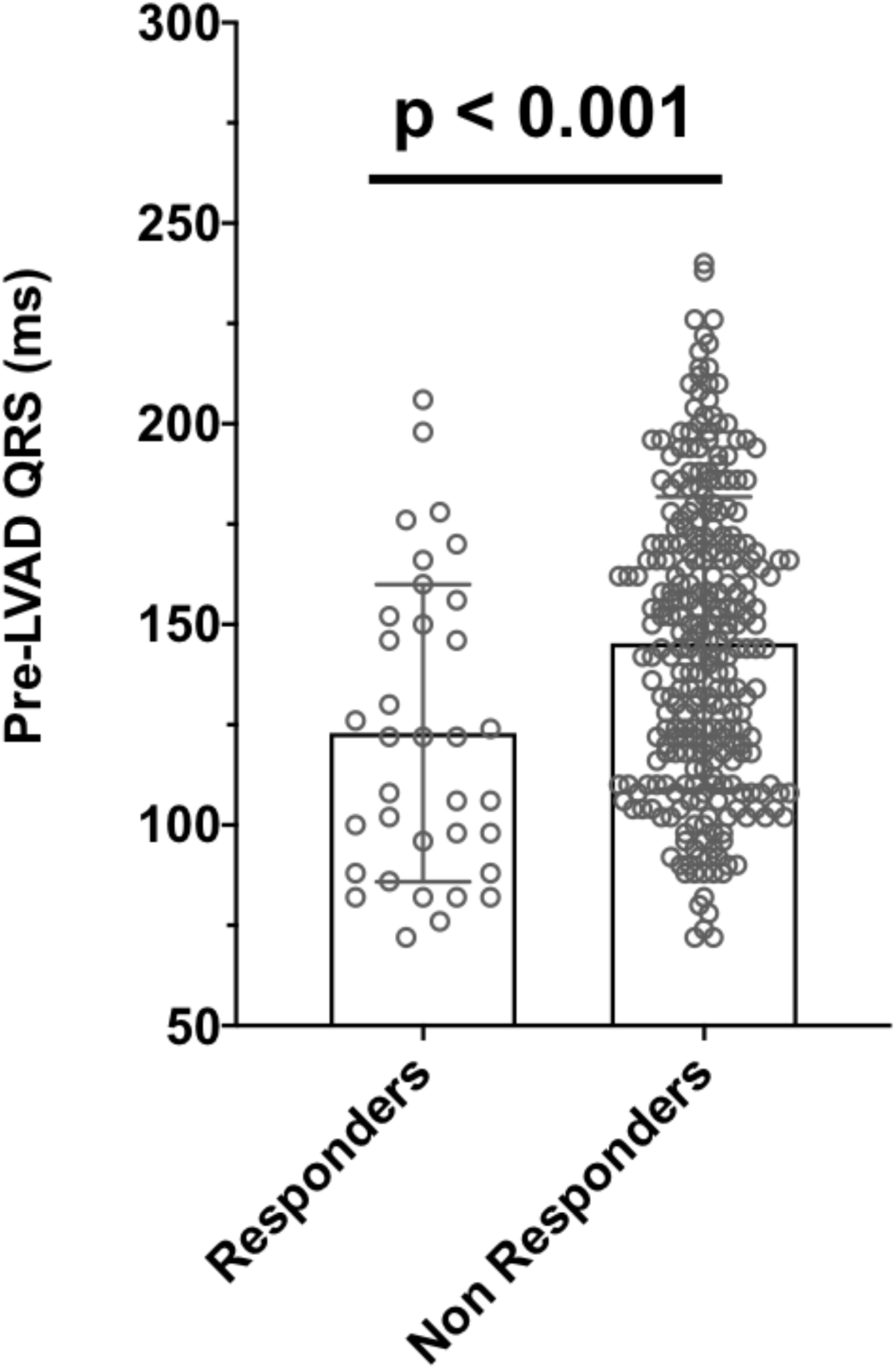
Baseline QRS duration in responders (n=35, 123±37 ms) in comparison to non-responders (n=280, 145±36 ms).

**Table 2.** Univariate logistic regression results of selected clinical parameters. Odds ratios (OR)<1.0 indicate the odds of cardiac recovery.

| Variables | OR | 95% CI | p value |
| --- | --- | --- | --- |
| Age | 0.968 | 0.947-0.989 | 0.003 |
| Male | 0.487 | 0.213-1.114 | 0.074 |
| BSA | 0.226 | 0.053-0.935 | 0.043 |
| BMI | 0.962 | 0.902-1.021 | 0.237 |
| Diabetes | 0.736 | 0.348-1.558 | 0.423 |
| Hypertension | 0.612 | 0.298-1.256 | 0.180 |
| NYHA Class IV | 0.699 | 0.337-1.448 | 0.335 |
| MCS | 1.846 | 0.499-6.818 | 0.358 |
| AF History | 0.507 | 0.225-1.067 | 0.084 |
| Previous Thoracotomy | 0.252 | 0.075-0.846 | 0.026 |
| Ischemic HF | 0.384 | 0.169-0.873 | 0.022 |
| Duration of HF | 0.988 | 0.981-0.995 | <0.001 |
| β-blockers | 1.633 | 0.739-3.611 | 0.226 |
| ACEI | 1.624 | 0.801-3.291 | 0.179 |
| Diuretics | 0.328 | 0.098-1.092 | 0.069 |
| Albumin | 0.488 | 0.237-1.000 | 0.051 |
| Bilirubin | 1.173 | 0.874-1.573 | 0.287 |
| ALP | 1.004 | 0.999-1.012 | 0.071 |
| Cr | 0.708 | 0.341-1.469 | 0.354 |
| BUN | 0.982 | 0.957-1.008 | 0.176 |
| K | 0.601 | 0.312-1.159 | 0.129 |
| BNP | 1.000 | 1.000-1.000 | 0.014 |
| HR | 1.016 | 0.998-1.034 | 0.072 |
| PAP | 0.975 | 0.941-1.010 | 0.165 |
| PVR | 0.916 | 0.774-1.083 | 0.305 |
| CI | 1.502 | 0.889-2.538 | 0.128 |
| LVEF | 0.992 | 0.942-1.045 | 0.776 |
| LVEDD | 0.663 | 0.449-0.985 | 0.042 |
| LVESD | 0.719 | 0.502-1.031 | 0.073 |
| IVSD | 0.377 | 0.083-1.720 | 0.208 |
| QRSd | 0.986 | 0.976-0.992 | <0.001 |

### Correlation of Baseline QRSd with Pre- and Post-LVAD LVEF and LVEDD in LVAD Patients

Before LVAD implant, as shown in **Fig. 3a**, there is a weak and non-significant correlation between QRSd and pre-LVAD LVEF in LVAD patients (r=-0.04, p=0.494). Unlike LVEF, pre-LVAD LVEDD exhibits a significant correlation with baseline QRSd as shown in **Fig. 3d** (r=0.24, p<0.01). After CF-LVAD support, the baseline QRSd shows a significant correlation with post-LVAD LVEF (r=-0.20, p<0.001) and similarly with pre- to post-LVAD LVEF change (r=-0.19, p<0.001), as shown in **Fig. 3b** and **Fig. 3c**, respectively. Nevertheless, baseline QRSd correlates non-significantly with post-LVAD LVEDD (r=0.07, p=0.208) and pre- to post-LVAD LVEDD change (r=-0.01, p=0.966) as shown in **Fig. 3e** and **2f**, respectively.

**Figure 3.**
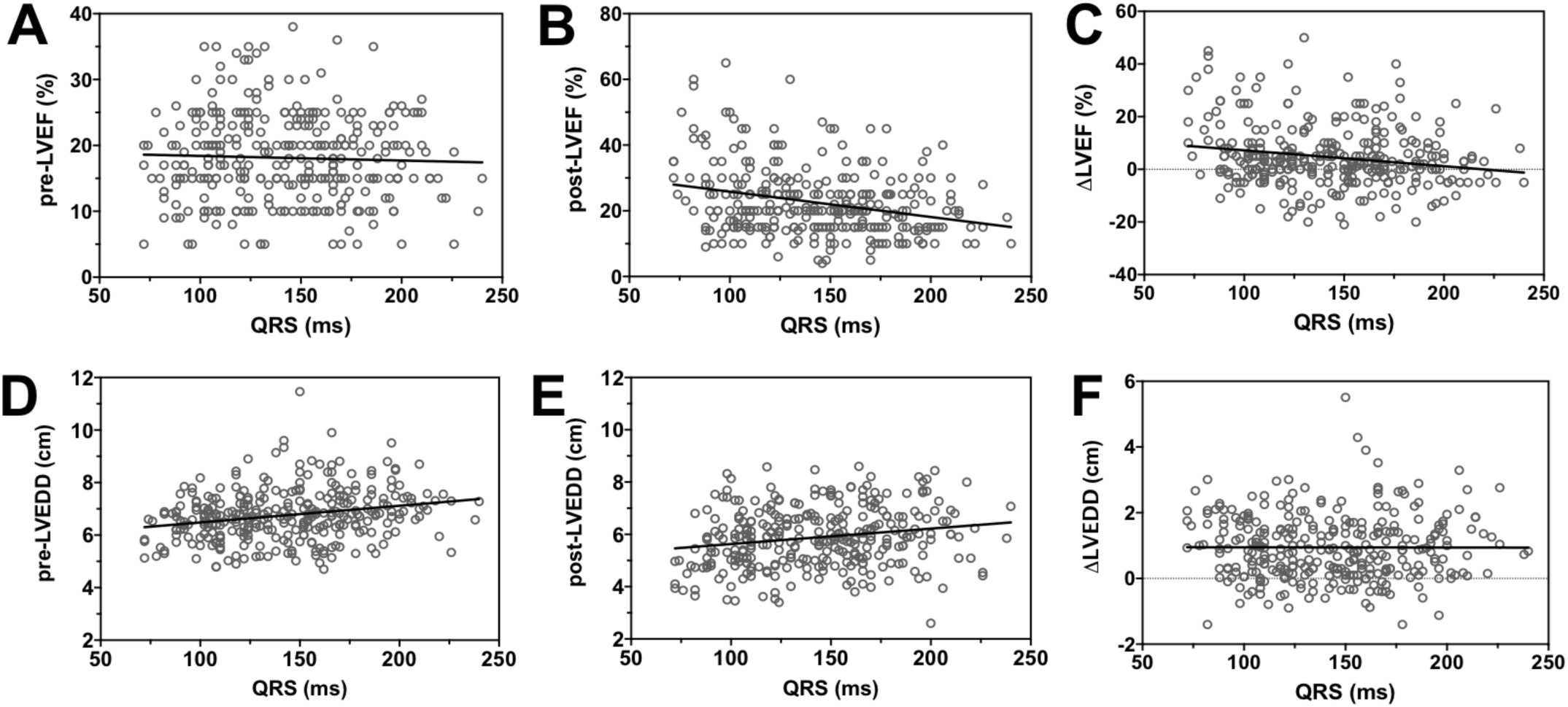
Impact of baseline QRS duration on pre- and post-LVAD LVEF (**Top Row**) and LVEDD (**Bottom Row**) in LVAD patients, n=315. **(A)** Correlation between QRS and pre-LVAD LVEF (r=-0.04, p=0.494). **(B)** Correlation between QRS and post-LVAD LVEF (r=-0.20, p<0.001). **(C)** Correlation between QRS and change (pre- to post-LVAD) in LVEF (r=-0.19, p<0.001). Here the negative ‘-’ in Pearson coefficient indicates the negative slope where EF reduces with increase in QRS duration. **(D)** Correlation between QRS and pre-LVAD LVEDD (r=0.24, p<0.001). **(E)** Correlation between QRS and post-LVAD LVEDD (r=0.07, p=0.208). **(F)** Correlation between QRS and change (pre- to post-LVAD) in LVEDD (r=-0.01, p=0.966).

### Correlation of Baseline QRSd with Pre- and Post-LVAD LVEF in Responders and Non-Responders

The relationship of baseline QRSd with LVEF before and after LVAD support, and pre- to post-LVAD change in LVEF in the ‘R’ group was investigated (**Top row**) as shown in **Fig. 4** and the results were compared with the ‘NR’ group (**Bottom row**). As shown in **Fig. 4a**, the ‘R’ group shows a significant improvement in LVEF (18±8 vs. 46±7 %, p<0.001) before and after LVAD implantation. However, the baseline QRSd is poorly and non-significantly correlated with pre-LVAD LVEF (r=0.15, p=0.386), post-LVAD LVEF (r=-0.15, p=0.386) and ΔLVEF (r=-0.22, p=0.205) as shown in **Fig. 4b, 3c**, and **3d**, respectively. Though after CF-LVAD support, the change in LVEF in ‘NR’ group is significant as shown in **Fig. 4e** (18±7 vs. 22±6 %, p<0.001), this group did not fulfill the criteria for post-LVAD cardiac recovery. Similarly, in the ‘NR’ group, the baseline QRSd is poorly and non-significantly related with pre-LVAD LVEF (r=-0.07, p=0.250), post-LVAD LVEF (r=-0.07, p=0.257) and ΔLVEF (r=-0.10, p=0.104) as shown in **Fig. 4f, 3g** and **3h**, respectively.

**Figure 4.**
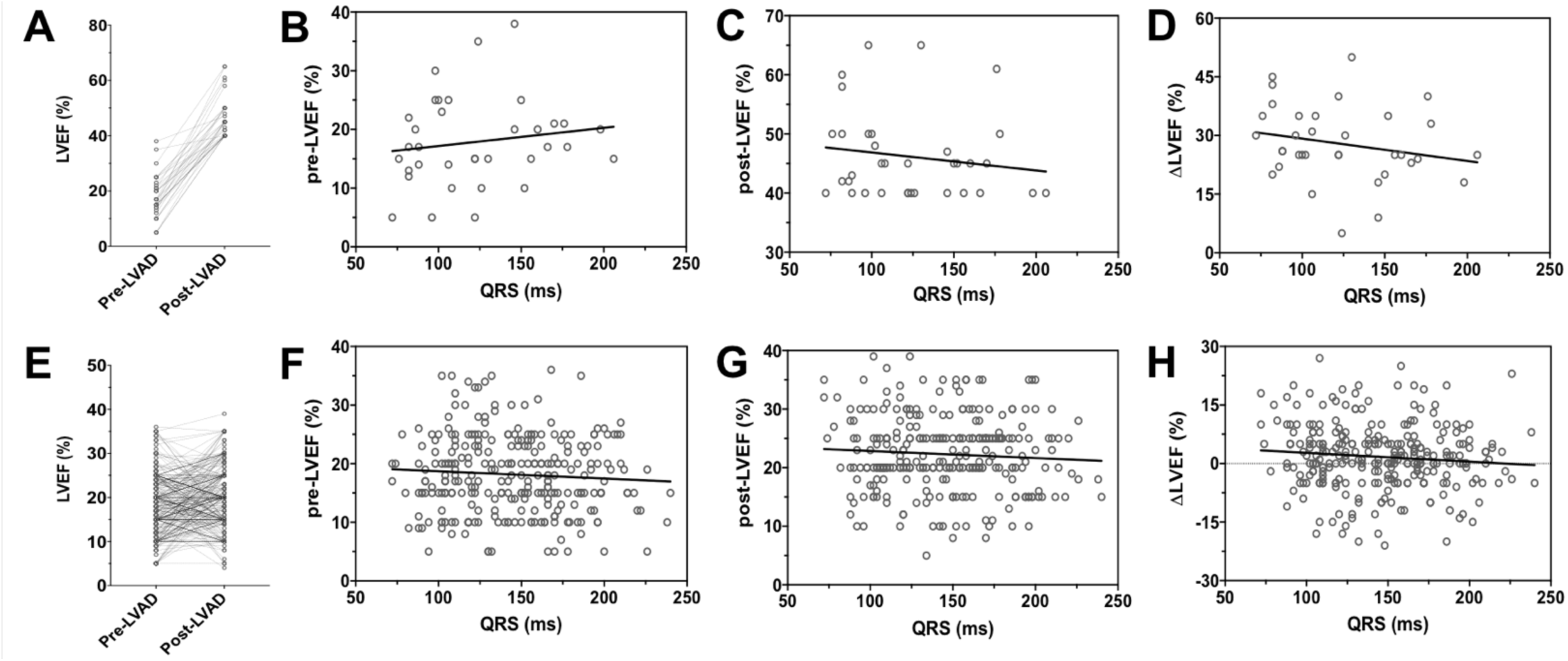
Impact of baseline QRS duration on pre- to post-LVAD LVEF in responders (n=35, **Top Row**) and non-responders (n=280, **Bottom Row**). **(A)** Comparing pre- and post-LVAD LVEF in responders (18±8 vs. 46±7 %, p<0.001). **(B)** Correlation between QRS and pre-LVAD LVEF (r=0.15, p=0.386). **(C)** Correlation between QRS and post-LVAD LVEF (r=-0.15, p=0.386). **(D)** Correlation between QRS and change (pre- to post-LVAD) in LVEF (r=-0.22, p=0.205). **(E)** Comparing pre- and post-LVAD LVEF in non-responders (18±7 vs. 22±6 %, p<0.001). **(F)** Correlation between QRS and pre-LVAD LVEF (r=-0.07, p=0.250). **(G)** Correlation between QRS and post-LVAD LVEF (r=-0.07, p=0.257). **(H)** Correlation between QRS and change (pre- to post-LVAD) in LVEF (r=-0.10, p=0.104). The negative sign ‘-’ in Pearson coefficient indicates the negative slope where change in LVEF reduces with increase in QRS duration.

### Correlation of Baseline QRSd with Pre- and Post-LVAD LVEDD Change in Responders and Non-Responders

The impact of baseline QRSd on pre-LVAD LVEDD, post-LVAD LVEDD and ΔLVEDD (pre- to post-LVAD change in LVEDD) in the ‘R’ group (**Top row**) was studied as shown in **Fig. 5**, and compared with ‘NR’ group (**Bottom row**). As shown in **Fig. 5a** and **Fig. 5e**, both groups (‘R’ and ‘NR’) show a significant improvement in post-LVAD LVEDD in comparison to their pre-LVAD LVEDD measures, respectively. Specifically, the ‘R’ group exhibits a 28% improvement in LVEDD following LVAD implant (6.4±0.9 vs. 4.6±0.6 cm, p<0.001). However, there was no correlation of the baseline QRSd with the pre- and post-LVAD LVEDD, as shown in **Fig. 5b** (r=0.19, p=0.268) and **Fig. 5c** (r=0.19, p=0.273), respectively. Similarly, in the ‘R’ group, the pre- to post-LVAD change in LVEDD is not correlated with baseline QRSd as shown in **Fig. 5d** (r=0.06, p=0.724). On the other side, in the ‘NR’ group, a 12% improvement in LVEDD is reported following LVAD implant (6.8±1.0 vs. 6.0±1.0 cm, p=p<0.001) as shown in **Fig. 5e**, whereas their baseline QRSd is poorly and non-significantly correlated with pre- to post-LVAD LVEDD change, as shown in **Fig. 5h** (r=-0.06, p=0.315).

**Figure 5.**
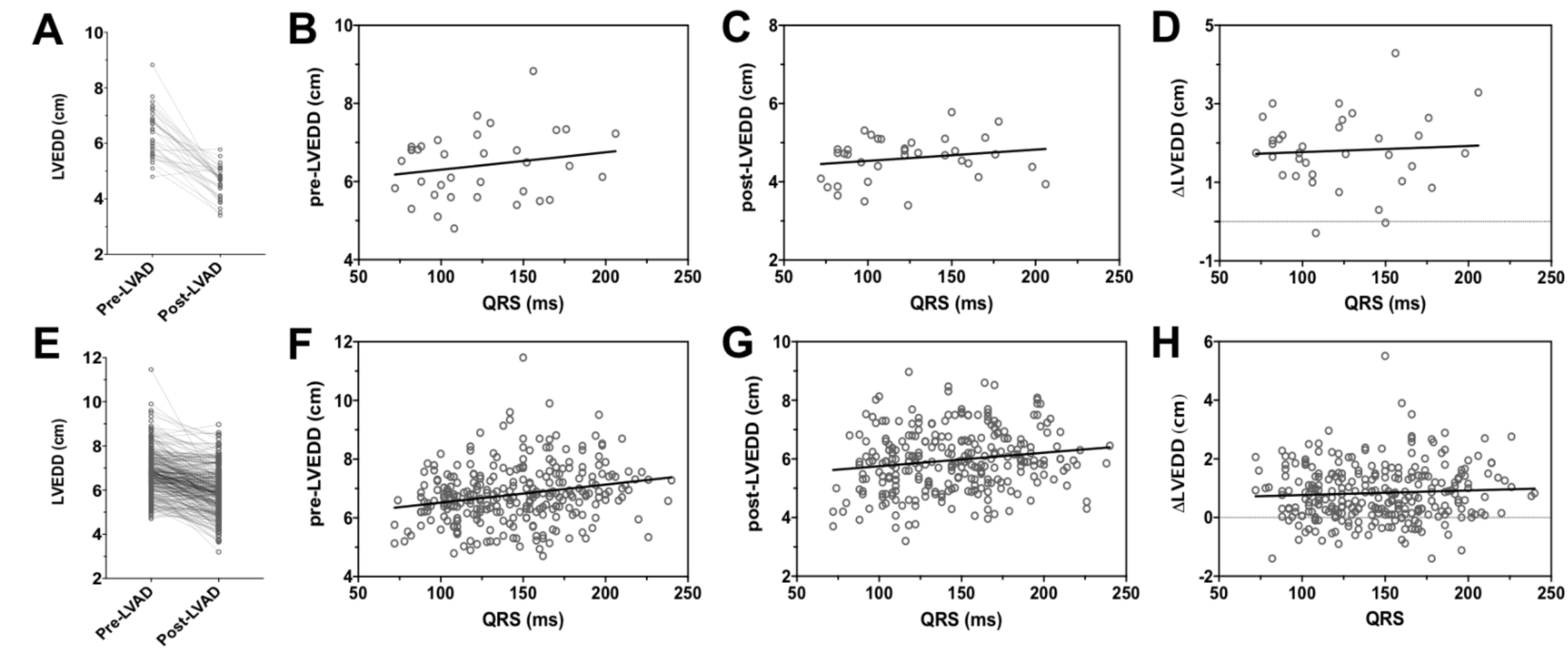
Impact of baseline QRS duration on pre- to post-LVAD LVEDD in responders (n=35, **Top Row**) and non-responders (n=280, **Bottom Row**). **(A)** Comparing pre- and post-LVAD LVEDD in responders (6.4±0.9 vs. 4.6±0.6 cm, p<0.001). **(B)** Correlation between QRS and pre-LVAD LVEDD (r=0.19, p=0.268). **(C)** Correlation between QRS and post-LVAD LVEDD (r=0.19, p=0.273). **(D)** Correlation between QRS duration and change (pre- to post-LVAD) in LVEDD (r=0.06, p=0.724). **(E)** Comparing pre- and post-LVAD LVEDD in non-responders (6.8±1.0 vs. 6.0±1.0 cm, p<0.001). **(F)** Correlation between QRS and pre-LVAD LVEDD (r=0.23, p<0.001). **(G)** Correlation between QRS and post-LVAD LVEDD (r=0.16, p=0.007). **(H)** Correlation between QRS and change (pre- to post-LVAD) in LVEDD (r=-0.06, p=0.315).

### Bivariate and Multivariate Analysis

In the bivariate models (**Table 3**), baseline QRSd is significantly associated with cardiac recovery after adjusting for age (OR: 0.983, 95% CI: 0.972-0.993, p=0.001), BSA (OR: 0.222, 95% CI: 0.049-0.946, p=0.046), previous thoracotomy (OR: 0.298, 95% CI: 0.069-0.878, p=0.053), ischemic HF etiology (OR: 0.430, 95% CI: 0.175-0.949, p=0.047, and BNP (OR: 1.000, 95% CI: 1.000-1.001, p=0.005). No other variables are associated with cardiac recovery in bivariate models when adjusted for baseline QRSd, though the baseline QRSd remained significant in those models (**Table 3)**.

**Table 3.** Bivariate logistic regression analysis to determine independent predictors of LVAD-induced cardiac recovery. OR<1.0 indicate the odds of cardiac recovery.

| Variables | OR | 95% CI | p value | AUC |
| --- | --- | --- | --- | --- |
| Age | 0.980 | 0.957-1.004 | 0.105 | 0.668 |
| QRSd | 0.987 | 0.975-0.998 | 0.022 |  |
| BSA | 0.222 | 0.049-0.946 | 0.046 | 0.671 |
| QRSd | 0.983 | 0.972-0.993 | 0.002 |  |
| Prev. Thoracotomy | 0.298 | 0.069-0.878 | 0.053 | 0.695 |
| QRSd | 0.984 | 0.973-0.995 | 0.004 |  |
| Ischemic HF | 0.430 | 0.175-0.949 | 0.047 | 0.684 |
| QRSd | 0.984 | 0.973-0.994 | 0.003 |  |
| Duration of HF | 0.990 | 0.982-0.996 | 0.006 | 0.726 |
| QRSd | 0.989 | 0.977-0.999 | 0.042 |  |
| Albumin | 0.498 | 0.236-1.032 | 0.063 | 0.681 |
| QRS | 0.983 | 0.972-0.993 | 0.002 |  |
| BNP | 1.000 | 1.000-1.001 | 0.005 | 0.701 |
| QRSd | 0.982 | 0.971-0.992 | 0.001 |  |
| LVEDD | 0.753 | 0.492-1.124 | 0.177 | 0.680 |
| QRSd | 0.985 | 0.973-0.995 | 0.005 |  |

The multivariate model (**Table 4**) with three parameters including baseline QRSd (OR: 0.987, 95% CI: 0.977-0.998, p=0.027), duration of HF (OR: 0.990, 95% CI: 0.983-0.997, p=0.006) and gender: male (OR: 0.388, 95% CI: 0.160-0.943, p=0.037) shows association for predicting post-LVAD cardiac recovery in LVAD patients with an accuracy of 0.73 (p<0.0001) as shown in **Fig. 6**.

**Table 4.**
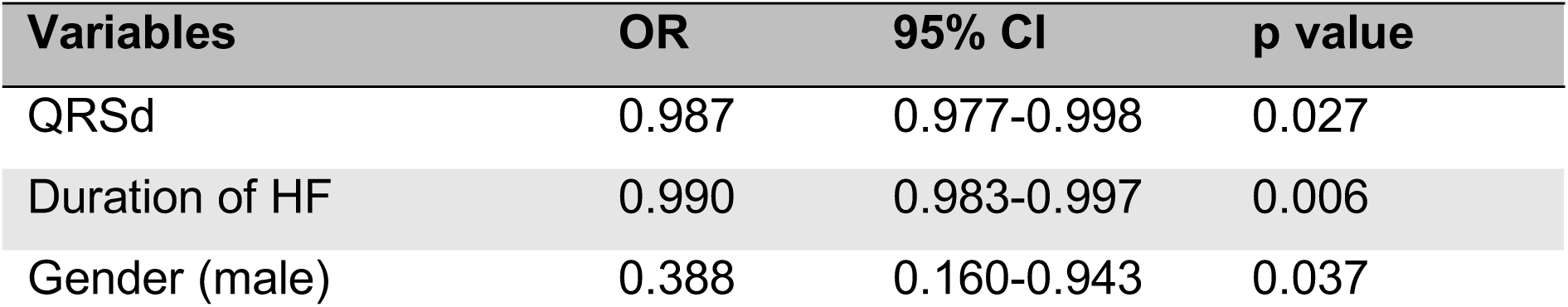
Multivariate logistic regression model with AUC: 0.73 (p<0.001).

**Figure 6.**
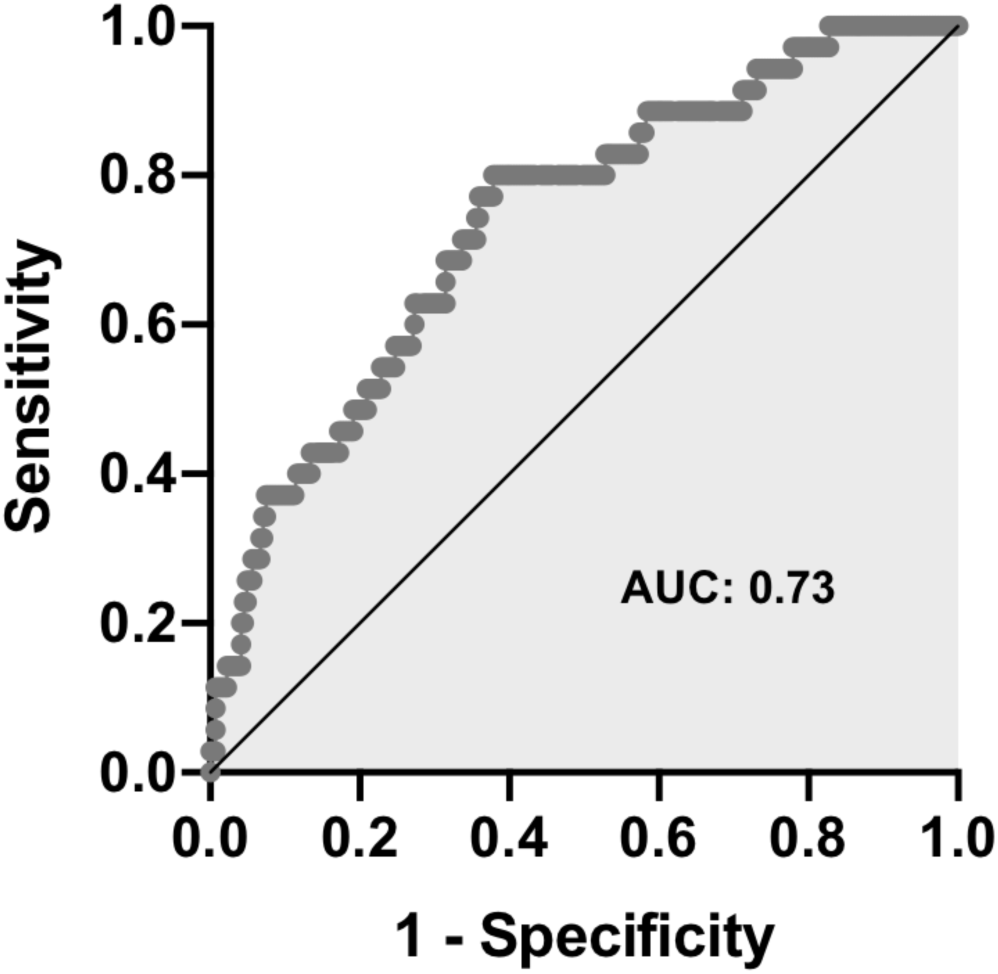
Multivariate logistic regression model comprised of baseline QRSd, HF duration and gender (male) shows an accuracy of 0.73 with p<0.001 predicting cardiac recovery within 12-months post-LVAD support.

## DISCUSSION

In chronic HF patients undergoing LVAD implantation, baseline QRSd was found to be associated with post-LVAD cardiac recovery within 12-months post-LVAD implantation. The pre-LVAD QRSd in the ‘R’ group was significantly shorter (15%) in comparison to the ‘NR’ group. It is noteworthy that patients who experienced cardiac recovery following LVAD implantation had a baseline LVEF similar to those who did not show post-LVAD cardiac recovery (**Table 1**). A comparable LVEF in the two groups is also consistent with previous studies.^8,18,24^

Previous studies have identified non-ischemic HF etiology, younger age and LV torsional mechanics as independent predictors of cardiac recovery.^9,25,26^ In concordance to these findings, our univariate data analysis showed that LVAD patients who achieved cardiac recovery within 12-months after LVAD implant were also more likely to be younger, female patients with a history of non-ischemic cardiomyopathy. Additionally, in responders the time from the HF diagnosis to the implantation of the LVAD was significantly shorter, a previous thoracotomy was less common, and there was a trend towards significance for higher baseline BNP levels compared to non-responders.

Previous studies have focused on a prolonged QRSd that appears common in patients suffering from HF with reduced LVEF.^14–16^ These studies emphasized that a baseline QRSd above ≥120 ms was associated with a significantly increased risk of death compared with a baseline QRSd <120 ms. Two prospective studies with 36 and 12 LVAD-supported patients, respectively, have previously investigated the QRS complex shortening at different time points during mechanical unloading. However, these studies did not explore an investigation of the effect of pre-LVAD QRSd on LVAD-induced cardiac recovery. Similarly, another prospective study of 23 LVAD patients, investigated the trajectory of QRSd immediately prior to LVAD implantation, and subsequently early and late while on LVAD support .^27^ Their findings did not focus on whether the pre-LVAD QRSd may predict the post-LVAD cardiac recovery in the same group of patients, instead they reported the comparison of baseline QRSd in LVAD patients (n=23) with another 22 control patients undergoing coronary artery bypass grafting. None of these studies reported the relationship of pre-LVAD QRSd in LVAD patients to cardiac recovery. One of the potential reasons could be their small sample size.

Based on a meta-analysis, the effect of very low LVEF and prolonged QRSd on the mortality benefits of ICD therapy has been reported in the general HF population.^30^ Further, pre- and post-LVAD fragmented QRS complex was studied in 98 LVAD patients to seek its association with survival following LVAD implantation over a 30-month follow-up period.^28^ Their results were based on the prevalence of fragmented QRS quantified at anterior, inferior and lateral territories. They did not distinguish the role of fragmented QRS as a predictor of cardiac recovery following LVAD support. The impact of baseline QRSd on pre- to post-LVAD change in LVEF has not been previously studied in CHF patients undergoing LVAD implantation, although the relation between fragmented QRS and LVEF has been discussed in HF patients.^29^ s

Similarly, the impact of baseline QRSd on LVEDD before and after LVAD support in chronic HF patients has not been reported yet. A positive association between QRSd and LV size in patients with bundle branch block was discussed previously,^31,32^ and it has been suggested that LV size does not modify the effect of baseline QRSd and its association with outcomes following cardiac resynchronization therapy.^33^ In our study, we observed a significant correlation of baseline QRSd with pre-LVAD LVEDD and a non-significant correlation with post-LVAD LVEDD as shown in **Fig. 3d** and **Fig. 2e**, respectively. At a first glance, this suggests that QRSd could play a vital role with pre-LVAD LVEDD in LVAD patients, however, the proposed scientific evidence is not true neither for the individuals who improved their cardiac structure and function while on LVAD support, (**Fig. 4b**) nor for those who did not (**Fig. 4f**). Compared to the ‘NR’ group that exhibited a 12% improvement in LVEDD, the ‘R’ group showed a 28% improvement from pre- to post-LVAD change in LVEDD, and these data indicate that QRSd may reflect the dimension and muscle mass of the LV and may be a useful indicator of LVAD-induced cardiac recovery.

## STUDY LIMITATIONS

The number of patients that fulfilled the criteria for post-LVAD cardiac recovery was relatively small (n=35). Future studies with a larger sample size are warranted to further explore the role of LV electrical remodeling in LVAD-induced cardiac recovery. LVAD patients with bundle branch block should ideally be studied separately regarding their baseline QRSd and the cardiac recovery potential. Integrating pre-LVAD QRSd to previously reported pre-LVAD clinical and translational cardiac recovery predictors may provide a highly sensitive and patient-specific electro-mechanistic method for predicting cardiac structural and functional improvement after LVAD unloading.

## CONCLUSIONS

Baseline QRSd effectively identified a subset of advanced cardiomyopathy patients prone to improve their cardiac structure and function following LVAD support. It could serve as a useful clinical indicator to guide the implementation of systematic monitoring and treatment strategies to promote cardiac recovery in selected LVAD candidates. Future research is warranted to further explore the association of baseline electrocardiographic indices with LV structural changes during mechanical support. Finally, strategies to facilitate cardiac recovery should be encouraged in such patients with the ultimate goal of LVAD weaning.

## Data Availability

The datasets generated during and/or analyzed during the current study are available from the corresponding author on reasonable request.

## Additional Information

Supplementary information is available for this paper in the supporting document attached to this file.

## Acronyms

(AF): Atrial fibrillation
(ALT): Alanine aminotransferase
(AST): Aspartate aminotransferase
(ALP): Alkaline phosphatase
(ACEI): Angiotensin-converting-enzyme inhibitor
(ARB): Angiotensin receptor blockers
(BUN): Blood urea nitrogen
(BNP): Brain natriuretic peptide
(BSA): Body surface area
(BMI): Body mass index
(CI): Cardiac index
(Cr): Creatinine
(HR): Heart rate
(Hb): Hemoglobin
(IABP): Intra-aortic balloon pump
(IVSD): Inter ventricular septal diameter
(LVEF): Left ventricular ejection fraction
(LVEDD): Left ventricular end-diastolic diameter
(LVESD): Left ventricular end-systolic diameter
(MCS): Mechanical circulatory support
(RAP): Right atrial pressure
(PAP): Pulmonary atrial pressure
(PCWP): Pulmonary capillary wedge pressure
(PVR): Pulmonary vascular resistance
(RVSWI): Right ventricular stroke work index
(SVR): Systemic vascular resistance
(TR): Tricuspid regurgitation

